# Seroprevalence of COVID-19 in HIV Population

**DOI:** 10.1101/2021.06.17.21259066

**Authors:** Shivdas Rajaram Naik, Swasthi S Kumar, Ankit Mittal, Satish Swain, Sanjay Ranjan, Manish Soneja, Sanjeev Sinha, Neeraj Nischal, Pankaj Jorwal, Pradeep Chaturvedi, Naveet Wig

**Affiliations:** Infectious Diseases, Department of Medicine, AIIMS, New Delhi, India; Department of Medicine, AIIMS, New Delhi, India; ART Centre, Department of Medicine, AIIMS, New Delhi, India; Department of Reproductive Biology, AIIMS, New Delhi, India

**Keywords:** HIV, Seroprevalence, PLHA, COVID-19, SARS-CoV-2 antibodies

## Abstract

**Background:** Seroprevalence helps us to estimate the exact prevalence of a disease in a population. Although the world has been battling this pandemic for more than a year now, we still do not know about the burden of this disease in people living with HIV/AIDS (PLHA). Seroprevalence data in this population subset is scarce in most parts of the world, including India. The current study aimed to estimate the seroprevalence of anti-SARS-CoV-2 IgG antibody among PLHA.

**Aim:** To determine the seroprevalence of SARS-CoV-2 antibodies in PLHA.

**Method:** This was a cross-sectional study conducted at a tertiary care hospital in North India. We recruited HIV positive patients following at the ART centre of the institute. Anti-SARS-CoV-2 IgG antibody levels targeting recombinant spike receptor-binding domain (RBD) protein of SARS CoV-2 were estimated in serum sample by the chemiluminescent immunoassay method.

**Results:** A total of 164 patients were recruited in the study with a mean age (±SD) of 41.2 (±15.4) years, of which 55% were male. Positive serology against SARS CoV-2 was detected in 14% patients (95% CI: 9.1-20.3%).

**Conclusion:** The seroprevalence of COVID-19 infection in PLHA was lower than the general population in the same region, which ranged from 23.48% to 28.3% around the study period.

## Introduction

Since the beginning of the COVID-19 pandemic, there is a fear that people living with HIV-AIDS (PLHA) are at increased risk of acquiring the disease. Also, they were presumably at risk of running a severe course of illness and have higher mortality. These assumptions were based on the observations made in other immunocompromised states (1). However, to date, no study has been able to show this association. On the contrary, recent studies have shown that these groups of people have a lower incidence of COVID-19 (2). However, a lot remains to be known about the HIV-COVID-19 relationship.

As the patients with COVID-19 are mostly asymptomatic, and developing countries are still unable to reach the desired number of RT PCR tests for diagnosis, surveillance for active cases becomes difficult. This is further compounded by the low sensitivity of the tests (3). In such situations, serological tests can often help us estimate the disease’s overall prevalence in a specified population.

As per a study conducted by Sharma et al., the adjusted seroprevalence of COVID-19 in Delhi during August, September, and October was 28.39%, 24.08%, and 24.71% (4). In another study conducted by the Indian Council of Medical Research (ICMR), the national seroprevalence was 0.7% during May-June and 7% during August-September 2020, with higher prevalence in urban areas (5,6).

We conducted this study to estimate the seroprevalence of COVID-19 amongst PLHA following up at the institutional anti-retroviral therapy (ART) centre.

## Methodology

This was a cross-sectional study designed to determine the seroprevalence of SARS-CoV-2 antibodies in PLHA. Participants were recruited from ART centre, All India Institute of Medical Sciences (AIIMS) New Delhi, between 1st September 2020 and 30th November 2020.

The Anti-Retroviral Therapy (ART) clinic works under the National AIDS Control Organization (NACO), an initiative of the Ministry of Health and Family Welfare of India’s Government. The hospital provides tertiary care to Delhi and the neighbouring States, and most of the patients come from the lower socio-economic strata. The centre provides testing for HIV, counselling (before and after testing), free of cost ART drugs and opportunistic infections (OI) screening and treatment.

PLHA visiting the centre, aged 18 years and above at the time of recruitment, were eligible to participate. Informed consent was obtained before the enrolment in the study. Patients presenting with features suggestive of active COVID-19 disease; were excluded.

The participants provided information at enrolment relating to age, sex, ART regime, symptoms suggestive of past COVID-19 infection or confirmed COVID-19 disease.

All the participants underwent phlebotomy performed by experienced medical staff. 2 ml Serum was drawn, and the samples were tested for antibodies to SARS-CoV-2using the Abbott Architect i4000SR (Abbott Diagnostics, Chicago, USA), which have been validated for use in adults (7).

The Abbott assay is highly specific for SARS-CoV-2 antibodies, using the manufacturer’s suggested cut-offs, with specificities of 1.00 (95% CI 0.98 to 1.00) and sensitivities at 0.94 (95% CI 0.86 to 0.98). The assay is a chemiluminescent immunoassay that detects IgG raised against the nucleocapsid protein of SARS-CoV-2. A signal/cut-off (S/CO) ratio of 1.4 was interpreted as reactive, and a S/CO ratio of less than 1.4 was considered non-reactive. Calibration was performed before initiating the test, and positive quality control (QC) S/CO 1.65-8.40 and negative quality control S/ CO < 0.78 were fulfilled before analyses of patient samples (7).

Study data were collected on a predesigned proforma. Primary outcome measures were to look for the presence of IgG antibodies to SARS-CoV-2 in serum. SARS-CoV-2 seropositivity was defined as a positive antibody test using the manufacturer’s advised positivity cut-off.

### Sample size justification

The study was powered to detect the seroprevalence of SARS-CoV-2 antibodies in PLHA. To achieve this, 163 participants were required (assuming an alpha of 0.05 and a beta of 0.2). At the beginning of the study, we expected an incidence of no more than 10% of COVID-19 in the HIV population. So, considering the prevalence in general population=20%, alpha error = 0.05, power= 95%, the sample size was calculated to be 163 (8).

### Statistical Analysis

Variables including sex, age, symptomatology, the regime of ART and SARS-CoV-2 antibody prevalence were analyzed using descriptive statistics (number and proportion for discrete variables, mean and SD for continuous variables). Variables associated with SARS-CoV-2 positivity were analyzed using univariate analyses to identify predictors of SARS-CoV-2 seropositivity. Initially, all possible variables were assessed using univariate analysis with Fisher’s exact testing of categorical data and the Mann-Whitney U test for continuous data. A p-value of less than 0.05 was considered statistically significant.

## Results

The study was conducted at the ART centre of AIIMS, New Delhi, between 1st September and 30th November 2020. During this period, 164 patients were recruited. 116 (70.1%) patients were males (table 1). The mean age (+SD) of the patients was 38.4 (+ 11.5) years. The median (IQR) CD4+Tcell count was 420.2 (268-543). There were 3 (1.8%) ART Naïve patients, 156 (95.1%) on first-line ART treatment (2 Nucleos(t)ide Reverse Transcriptase Inhibitor [NRTI]+ 1 Non-Nucleoside Reverse Transcriptase Inhibitor [NNRTI]) and 5 (3.1%) on second-line ART treatment (2 NRTI+ 1 Protease Inhibitor [PI]) (Table 2).

**Table 1:** Baseline details of the Patients.

|  | Total<br>(n=164) | Positive<br>(n=23) | Negative<br>(n=141) | P-value |
| --- | --- | --- | --- | --- |
| Male | 116 (70.7%) | 19 (82.6%) | 97 (68.8%) | 0.22 |
| Age (mean ± SD) | 38.4 ± 11.5 | 40.1 ± 11.7 | 38.0 ± 11.4 | 0.42 |
| Current cd4 count<br>(n=162) (median (IQR)) | 420.2<br>(268.5-543.75) | 335 (275-463.5) | 420<br>(267.5-546.5) | 0.30 |
| CD4+Tcell count |  |  |  | 0.37 |
| • Less than 200 cells/mm<sup>3</sup><br>• 200-349 cells/mm<sup>3</sup><br>• 350-499 cells/mm<sup>3</sup><br>• More than 500 cells/mm<sup>3</sup> | 17 (10.4%)<br>54 (32.9%)<br>38 (23.1%)<br>53 (32.3%) | 3 (13.4%)<br>11 (47.2%)<br>4 (17.4%)<br>5 (21.7%) | 14 (10%)<br>43 (30.5%)<br>34 (24.1%)<br>48 (34.0%) |  |

**Table 2:** ART regime of the patients recruited.

|  | Total [n (%)] | Positive<br>(n=23) | Negative<br>(n=141) |
| --- | --- | --- | --- |
| **<u>ART regime</u>** |  |  |  |
| Naïve | 3 (1.8) | 0 (0) | 3 (2.1) |
| First-line [2 NRTI + 1 NNRTI] | 156 (95.1) | 23 (100) | 133 (94.4) |
| Second Line [2NRTI + 1PI] | 5 (3.0) | 0 (0) | 5 (3.5) |
| **<u>Drug</u>** |  |  |  |
| TLE | 92 (56.1) | 18(78.3) | 74(52.5) |
| TLN | 3 (1.8) | 1(4.3) | 2(1.4) |
| ZLN | 53 (32.3) | 4(17.4) | 49(34.8) |
| ZLE | 7 (4.3) | 0(0) | 7(5.0) |
| TLA/r | 2 (1.2) | 0(0) | 2(1.4) |
| ZLA/r | 3 (1.8%) | 0(0) | 3(2.1) |

**T-tenofovir, L-lamivudine, E-efavirenz, N- Nevirapine, Z- zidovudine, A/r- ritonavir boosted atazanavir**

Twenty-Three (14%) patients were found to be positive for COVID-19 antibody by chemiluminescent immunoassay on the Abbott Architect i4000SR. Of this, 19 (82.6%) were males. This makes the overall prevalence of COVID-19 in the HIV population in the Delhi NCR region 14% (95%CI: 9.1-20.3%). Although seropositivity was higher in males (16.3% vs 8.3%), this difference was not statistically significant. Seroprevalence of COVID-19 was highest in PLHA aged more than 50 years (24%). No statistically significant difference among antibody positivity rate was noted in any age group. The median (IQR) CD4+Tcell count was 335 (275-463.5) and was lower than the COVID-19 antibody-negative group. But the difference was not statistically significant.

Also, 19 patients reported at least one symptom that could be suggestive of a previous COVID-19 infection. Of them, 12 (63.2%) had a fever, 10 (52.6%) had a cough, 11(57.9%) had a sore throat, and 2 (10.5%) patients had myalgia, rash and running nose. Out of these 19 patients, only 8 patients were tested for COVID-19 infection by RT PCR test. All tested negative. But, 4 of them were found to have antibodies against COVID-19.

## Discussion

This was the first study to look at the seroprevalence of COVID-19 infection in the HIV population in India. The seroprevalence of COVID-19 in PLHA in the Delhi NCR region was found to be 14%.

The serosurvey studies conducted by the Ministry of Health and Family Welfare in Delhi during June-July, August and October respectively revealed the seroprevalence of COVID-19 infection in the Delhi population to be 23.48%, 28.3% and 25.9% (9,10). A study conducted by Ray et al. in hospitalized patients revealed the seroprevalence to be 19.8% (11). When compared to the general population, the present study had a lower seroprevalence in the PLHA group. Similar results were seen in a survey conducted by Papalini et al. Their study, conducted during the early phase of the pandemic, also had a lower seroprevalence of COVID-19 in PLHA than the general population (12). This may be attributed to the fact that most of the patients were indoors, avoiding social contact due to fear of acquiring the disease and may not have contracted the disease (13). Another reason for this could be that these patients might not have generated antibodies against COVID-19 infection or may not have sustained it after getting infected. This is based on our knowledge of impaired immune response in PLHA (13,14). The role of a few ART drugs, especially lopinavir-ritonavir, was considered a potential for the treatment of COVID-19 in the initial days (15). Subsequent literature found it not to be effective (16). However, the role of other anti-retroviral drugs or their combination is unknown and unexplored.

The majority of the patients in the study had no or minimal symptoms of COVID-19 infection. Since the symptoms are non-specific, they cannot always be attributed to COVID-19 infection. In South Korea, more than 60% of the PLHA patients were asymptomatic for COVID-19 at the time of diagnosis (17). We also know that at least 40-45% of patients will remain asymptomatic for COVID-19 (18). Another study by Ray et al. at the institute had revealed that most of the patients (>97%) had minor or no symptoms. This likely suggests that, like the general population, asymptomatic infections are also prevalent in the PLHA.

There were limitations to the study. As it was a single centre-based study, it may not be an accurate representation of this subgroup. There were a smaller number of patients who were naïve or on ART regimens other than TLE (tenofovir, lamivudine, efavirenz) and ZLN (zidovudine, lamivudine, nevirapine) combination. Also, since anti-SARS-CoV-2 antibody levels are known to wane over a few months, some cases might have been inadvertently missed in this cross-sectional study (19). Also, the possibility of false results, as seen with any other antibody-based test, cannot be ruled out.

## Conclusion

The present study indicates the seroprevalence of COVID-19 infection in PLHA is lower than the general population. However, the exact reasons explaining this lower seroprevalence is still not clear. Furthermore, this seroprevalence is bound to change with time and subsequent waves of COVID-19 infection and therefore, there is a need for continuous serosurveillance to understand the infection dynamics in this subpopulation.

## Data Availability

Data is saved with the first author and can be made available if requested. but this is not stored on any URLs

## Author contributions

Shivdas Naik, Ankit Mittal, Manish Soneja conceived the idea. Shivdas Naik and Ankit Mittal prepared the first draft. Manish Soneja, Sanjeev Sinha, Neeraj Nischal, Pankaj Jorwal, and Naveet Wig helped review the literature. Shivdas Naik, Swasthi Kumar, Satish Swain, Sanjay Ranjan were involved in sample collection and data acquisition. Pradeep Chaturvedi was involved in sample processing and antibody testing. Shivdas Naik and Manish Soneja were involved in data analysis and interpretation. Manish Soneja, Sanjeev Sinha, Neeraj Nischal, Pankaj Jorwal, Pradeep Chaturvedi and Naveet Wig did the proofreading of the article. Finally, all authors reviewed the manuscript.

## Conflicts of interest

We declare no conflicts of interest.

## Ethical clearance

The study was cleared by the institutional ethics committee of AIIMS, New Delhi (IECPG/439/8/2020)

## Acknowledgement

We thank the staff at ART centre, AIIMS New Delhi, for their help and support in the study.

## Funding

No external funding.

## Notes

### Competing Interest Statement

The authors have declared no competing interest.

### Author Declarations

The study was cleared by the institutional ethics committee of AIIMS, New Delhi (IECPG/439/8/2020)

## References

1. Xu Z, Zhang C, Wang F-S. COVID-19 in people with HIV. Lancet HIV. 2020 Aug;7(8):e524–6.

2. del Amo J, Polo R, Moreno S, Díaz A, Martínez E, Arribas JR, Jarrin I and Hernán MA Incidence and Severity of COVID-19 in HIV-Positive Persons Receiving Antiretroviral Therapy: A Cohort Study. Ann Intern Med. 2020 Oct 6;173(7):536–41.

3. Watson J, Whiting PF, Brush JE. Interpreting a covid-19 test result. BMJ. 2020 May 12;m1808.

4. Sharma N, Sharma P, Basu S, Saxena S, Chawla R, Dushyant K, Mundeja N, Marak ZS, Singh S, Singh GK, Rustagi R. The seroprevalence and trends of SARS-CoV-2 in Delhi, India: A repeated population-based seroepidemiological study. medRxiv. 2020 Jan 1.

5. Murhekar MV, Bhatnagar T, Selvaraju S, Rade K, Saravanakumar V, Thangaraj JW, Kumar MS, Shah N, Sabarinathan R, Turuk A, Anand PK. Prevalence of SARS-CoV-2 infection in India: Findings from the national serosurvey, May-June 2020. Indian Journal of Medical Research. 2020 Jan 1;152(1):48.

6. Murhekar MV, Bhatnagar T, Selvaraju S, Saravanakumar V, Thangaraj JW, Shah N, Kumar MS, Rade K, Sabarinathan R, Asthana S, Balachandar R. SARS-CoV-2 antibody seroprevalence in India, August–September 2020: findings from the second nationwide household serosurvey. The Lancet Global Health. 2021 Mar 1;9(3):e257–66.

7. Bryan A, Pepper G, Wener MH, Fink SL, Morishima C, Chaudhary A, Jerome KR, Mathias PC, Greninger AL. Performance characteristics of the Abbott Architect SARS-CoV-2 IgG assay and seroprevalence in Boise, Idaho. Journal of clinical microbiology. 2020 Jul 23;58(8):e00941–20.

8. Rosner B. Fundamentals of biostatistics. 8th edition. Boston, MA: Cengage Learning; 2016. 927 p.

9. Seroprevalence study conducted by National Center for Disease Control NCDC, MoHFW, in Delhi, June 2020 [Internet]. [Cited 2020 Dec 23]. Available from: pib.gov.in/Pressreleaseshare.aspx?PRID=1640137

10. Seroprevalence almost 50% in Central district in Delhi | Cities News, The Indian Express [Internet]. [Cited 2020 Dec 23]. Available from: https://indianexpress.com/article/cities/delhi/seroprevalence-almost-50-in-central-district-in-delhi-7048490/

11. Ray A, Singh K, Chattopadhyay S, Mehdi F, Batra G, Gupta A, Agarwal A, Bhavesh M, Sahni S, Chaithra R, Agarwal S. Seroprevalence of anti-SARS-CoV-2 IgG antibodies in hospitalized patients at a tertiary referral centre in North India. medRxiv. 2020 Jan 1.

12. Papalini C, Paciosi F, Schiaroli E, Pierucci S, Busti C, Bozza S, Mencacci A, Francisci D. Seroprevalence of anti-SARS-CoV2 Antibodies in Umbrian Persons Living with HIV. Mediterranean Journal of Hematology and Infectious Diseases. 2020;12(1).

13. Karim F, Gazy I, Cele S, Zungu Y, Krause R, Bernstein M, Ganga Y, Rodel H, Mthabela N, Mazibuko M, Khan K. HIV infection alters SARS-CoV-2 responsive immune parameters but not clinical outcomes in COVID-19 disease. medRxiv. 2020 Jan 1.

14. Kerneis S, Launay O, Turbelin C, Batteux F, Hanslik T, Boelle P-Y. Long-term Immune Responses to Vaccination in HIV-Infected Patients: A Systematic Review and Meta-Analysis. Clin Infect Dis. 2014 Apr 15;58(8):1130–9.

15. Braus M, Morton B. Art therapy in the time of COVID-19. Psychol Trauma Theory Res Pract Policy. 2020 Aug;12(S1): S267–8.

16. WHO Solidarity Trial Consortium. Repurposed Antiviral Drugs for Covid-19 — Interim WHO Solidarity Trial Results. N Engl J Med. 2020 Dec 2;NEJMoa2023184.

17. Jung C-Y, Park H, Kim DW, Choi YJ, Kim SW, Chang TI. Clinical Characteristics of Asymptomatic Patients with COVID-19: A Nationwide Cohort Study in South Korea. Int J Infect Dis. 2020 Oct;99:266–8.

18. Oran DP, Topol EJ. Prevalence of Asymptomatic SARS-CoV-2 Infection: A Narrative Review. Ann Intern Med. 2020 Sep 1;173(5):362–7.

19. Choe PG, Kang CK, Suh HJ, Jung J, Song KH, Bang JH, Kim ES, Kim HB, Park SW, Kim NJ, Park WB. Waning antibody responses in asymptomatic and symptomatic SARS-CoV-2 infection. Emerging infectious diseases. 2021 Jan;27(1):327.

